# Factors influencing referral to maternity models of care in Australian general practice

**DOI:** 10.1101/2023.12.16.23300085

**Authors:** J Thomas, L Kuliukas, J Frayne, Z Bradfield

**Affiliations:** School of Nursing, Curtin University, Western Australia, Australia; Medical School, University of Western Australia, Western Australia, Australia

## Abstract

**Background:** In the Australian maternity system, general practitioners play a vital role in advising and directing prospective parents to maternity models of care. Optimising model of care discussions and the decision-making process avoids misaligning women with over or under specialised care, reduces the potential for disruptive care transitions and unnecessary healthcare costs, and is critical in ensuring consumer satisfaction. Current literature overwhelmingly focusses on women’s decision-making around model of care discussions and neglects the gatekeeping role of the General Practitioner (GP). This study aimed to explore and describe the factors influencing Australian GPs decision-making when referring pregnant women to maternity models of care.

**Methods:** This study used a qualitative descriptive approach. General practitioners (N=12) with experience referring women to maternity models of care in Australia participated in a semi-structured interview. Interviews occurred between October and November 2021 by telephone or videoconference. Reflexive thematic analysis was facilitated by NVivo-12 data management software to codify and interpret themes from the data.

**Findings:** Two broad themes were interpreted from the data. The first theme entitled ‘GP Factors’, incorporated three associated sub-themes including ‘GPs Previous Model of Care Experience’, ‘Gaps in GP Knowledge’ and ‘GP Perception of Models of Care’. The second theme, entitled ‘Woman’s Factors’, encapsulated two associated sub-themes including the ‘Woman’s Preferences’ and ‘Access to Models’.

**Conclusions:** This study provides novel evidence regarding general practitioner perspectives of the factors influencing model of care decision-making and referral. The exploration and description of factors influencing model of care decisions provide unique insight into the ways that all stakeholders can experience access to a broader range of models of care including midwifery-led continuity of care models aligned with consumer-demand. In addition, the role of national primary health networks is outlined as a means to achieving this.

## Introduction

The Australian healthcare system and its maternity services operate through a two-tier system of both public and private providers [1, 2]. The national publicly funded universal healthcare program, Medicare, provides hospital and community-based care to Australian and New Zealand citizens with predominantly no out-of-pocket costs [2, 3]. Conversely, private hospitals and services are accessible with private health insurance and a co-payment or alternatively, incur a full fee for service [2, 3].

Within this system, almost all pregnant women will make initial contact with a general practitioner (GP) when commencing their maternity care journey [4, 5]. GPs provide healthcare across the lifespan and play a key role in advising and directing prospective parents to a maternity model of care [2]. They have a gatekeeping function in the way that access to certain models of care is only granted through the GP referral. Although other referral pathways exist such as through other clinicians or via self-referral, the GP referral pathway is the most common way women access models of care [2, 6].

In Australia, there are 11 models of maternity care according to the Maternity Care Classification System (MaCCs), each has their own specific features, personnel, and fee structure [2, 7]. These can be grouped into five major model of care categories which include standard public care, shared care, public midwifery continuity care, private obstetric care and private midwifery care [2, 7]. Standard public care is provided by hospital doctors and midwives and is government funded [2, 7]. Shared care involves antenatal care delivered by a community doctor and or midwife, with several key antenatal visits and intrapartum care occurring in the public hospital [2, 7]. Hospital-based care within a shared care model is government funded though women may incur out of pocket costs for GP-led visits [2, 7]. Public midwifery continuity of care models are government funded and provided by a hospital midwife or small team of midwives with birth largely occurring in a birth centre or public hospital [2, 7]. Within the public midwifery continuity of care category is midwifery group practice caseload care. Midwifery group practice caseload care involves antenatal care provided by a known midwife or group of midwives in the hospital, community, home, or birth centre setting with intrapartum care taking place in the hospital, birth centre or home [2, 7]. Private obstetric care is provided by an obstetrician with birth occurring in a private hospital and is largely accessible via women’s private health insurance though some Medicare rebates may be available [2, 7]. Subsumed within the private obstetric care major model category is general practitioner obstetrician care which involves community or hospital clinic antenatal care and public or private hospital intrapartum care provided by a GP obstetrician [2, 7]. Finally, private midwifery care involves care by a privately practising midwife who supports birth in the home or hospital settings and can be partly government funded and partly consumer funded [2, 7].

It is known that 97% of Australian women intend to and will give birth in a hospital setting but it is not known how women distribute across the models of care at a national level as this is not yet reported [2, 8]. The first national model of maternity care report was published in November 2021, has been re-released annually and focusses on the maternity model of care characteristics [1, 8, 9]. As such, there remains limited data comparing and evaluating the models of care in Australia [2, 10–12]. Despite this, model of care report data and birthplace data can be used to infer how women distribute across the models of care. In 2023, approximately 1000 model of care services were active in Australia and most of these identified as public hospital maternity care (41%), 14% were midwifery group caseload care models, 15% identified as shared care models and 11% were private obstetrician specialist care models [8]. In 2019, Australian birthplace data revealed that 75% of women gave birth in a public hospital, 22% in a private hospital, 2.3% in a birth centre, 0.6% of women constituted births before the arrival to a health service and 0.3% of births occurred in the home setting [13].

## Significance

In 2009, the Australian Department of Health and Ageing published a maternity service review and recommended that there be improved model of care choice for women, a greater range of models of care be made available, an expanded role for midwives, improved access for Indigenous and rural women and further development of collaborative models of care [14]. This was strengthened by the most recent national maternity strategy built on four pillars of choice, safety, access and respect orientated by woman-centred care [15].

Model of care information, when comprehensive, allows parents to make trade-offs between the perceived advantages and disadvantages and select a framework that most aligns with their preferences [12, 16]. In addition, model of care decision making when optimised, enhances consumer satisfaction, prevents over or under specialised care, prevents excessive healthcare costs and the potential for disruptive care transitions [2, 17].

The Fourth National Atlas in Healthcare Variation which identifies potentially unwarranted variation in healthcare has highlighted early planned birth by caesarean section and induction of labour before 39 weeks without medical indication as an area of concern [18]. Early planned birth has been identified as a key contributor to neonatal respiratory problems, admission to neonatal intensive care units and longer-term cognitive deficits in children [18]. It is well accepted that midwifery-led continuity of care confers a range of benefits to mothers and infants including a higher chance of a normal labour and birth and less chance of intrapartum caesarean section [19, 20]. Specifically, women planning a homebirth have a six times greater chance of a vaginal birth compared to women planning a hospital birth and women planning a birth centre birth have a nearly double chance of vaginal birth compared to women planning a hospital birth without a continuity of care model [19, 20]. Moreover, midwifery continuity of care is the only health system intervention shown to reduce preterm birth and improve perinatal survival [19, 20]. Despite the range of consumer and health system benefits evident among midwifery-led continuity of care models, most Australian women are accessing public hospital care and it is the GP referral that often determines model of care and birthplace.

Models of care are also vital for the provision of culturally sensitive care for Aboriginal and Torres Strait Islander women and those from culturally and linguistically diverse (CALD) backgrounds. Midwifery-led continuity of care models that are tailored to the needs of Aboriginal and Torres Strait Islander women, particularly those that offer ‘birthing on country’, have been shown to improve health outcomes and confer a range of benefits to its consumers [21–25]. These models are unique in that they provide antenatal care alongside access to a range of multidisciplinary providers and are supported by Aboriginal Health Workers [22–24]. Improvements associated with these models include high levels of consumer satisfaction [22–24], improved antenatal attendance rates [25], improved breastfeeding rates [25], and a reduction in preterm birth rates [22, 26]. For CALD women, there are a range of service elements such as continuity of care, effective communication, culturally responsive care, support navigating systems and flexibility within services that align with maternal preferences, reduce barriers to access and have the potential to improve outcomes for CALD mothers and infants [27].

It is recognised that challenges navigating the referral system can act as a barrier to initiating antenatal care which is vital for the health and wellbeing of women and babies [28]. Finally, the literature overwhelmingly focusses on women’s decision-making and neglects the gatekeeping role of the GP [29–31]. Given the established importance of GP referral to model of care and dearth of evidence regarding factors that influence referral, this study aimed to explore and describe the factors influencing Australian GPs’ decision-making when referring pregnant women to maternity models of care.

## Methods

A qualitative description design was used given the recognised utility to derive rich descriptions from participants and its suitability for understanding novel phenomena/on [32, 33]. The research aim prompted the use of a critical realist lens which is ontologically realist asserting that an authentic reality exists separately to human knowing while also being epistemologically subjectivist asserting that multiple versions of reality can be produced [34]. A purposive, snowball and convenience sampling approach was taken utilising social media and email to recruit general practitioners from around the nation experienced in referring pregnant women to maternity models of care. Recruitment occurred from the 23^rd^ of August 2021 to the 20^th^ of October 2021. Prospective participants were provided with information on the study, the researchers involved, and its purpose; written consent was provided. GPs participated in a semi-structured interview conducted by researcher JT between October and November of 2021. Sampling was driven by data saturation, defined as the point at which limited new concepts or descriptions directly relevant to the research aim could be interpreted. Twelve general practitioners comprised the final sample size. Interviews were conducted via Microsoft Teams or via telephone and lasted approximately 30 minutes each. Demographic data were collected at the beginning of the interview. GP participants were asked “*Please describe how you decide where and to whom you refer your maternity clients?*”. Interviews were audio recorded and transcribed verbatim by researcher JT. Interview transcripts then underwent Braun and Clarke’s reflexive thematic analysis facilitated by NVivo-12 data management software including 1) familiarisation with the data, 2) coding the data, 3) identifying themes, 4) reviewing themes, 5) defining and naming themes, and 6) producing the report [35]. To enhance the rigor of the study, the researcher maintained an audit trail and engaged in peer-review at relevant intervals with the research team to expose bias or inappropriate subjectivity.

The data were managed utilising an encrypted, password protected cloud storage platform (OneDrive) and university research drive. Following the interviews, data copies were deleted from OneDrive and will remain on the university research drive for a period of seven years.

This study obtained ethical clearance from the Curtin University Human Research Ethics Committee in alignment with the National Health and Medical Research Council guidelines (HREC 2021-0500).

## Findings

The demographic profile of the participants is outlined in Table 1. Of the twelve participants, eleven identified as female. Participants were between 30 and 59 years of age with most participants aged between 40 and 49 years. Most participants were GP Fellows of the Royal Australian and New Zealand College of General Practitioners. Five participants had less than 10 years of experience working in general practice, five participants had between 11 and 20 years of experience and two participants had more than 20 years of experience working in general practice. Most participants were currently living and working in Western Australia. Most of the participating GPs were working in metropolitan areas however five participants were currently working in a rural or regional area.

**Table 1.** Demographic Profile of Participants.

| Demographic variables | Participant numbers<br>n=12 |
| --- | --- |
| **Gender** |  |
| Female | 11 |
| Male | 1 |
| **Age** |  |
| 30 to 39 | 4 |
| 40 to 49 | 6 |
| 50 to 59 | 2 |
| **Level of vocation** |  |
| GP Registrar | 1 |
| Fellow | 11 |
| **Years of experience as a GP** |  |
| 0 to 10 | 5 |
| 11 to 20 | 5 |
| 21 to 30 | 2 |
| **Additional obstetric qualification** |  |
| Nil | 5 |
| Basic Diploma | 6 |
| Advanced Diploma | 1 |
| **Maternity model work experience** |  |
| Nil | 2 |
| Birth Centre | 3 |
| GP Obstetric | 7 |
| Private hybrid model | 1 |
| Hospital | 10 |
| **Years of experience in maternity model** |  |
| Nil | 2 |
| < 1 year | 4 |
| 1 to 5 years | 3 |
| 6 to 10 years | 1 |
| Over 10 years | 2 |
| **Geographical classification** |  |
| Metropolitan | 7 |
| Rural | 4 |
| Regional | 1 |
| **State** |  |
| WA | 8 |
| NSW | 3 |
| QLD | 1 |

Two main themes were generated from the data analysis process. The first theme, entitled “GP factors” incorporates three subthemes including “Gaps in GP knowledge”, “GPs previous model of care experience” and “GPs perception of the models of care”. The second theme or the “Woman’s factors” encapsulates two subthemes including “Woman’s preferences” and “Access to models” (Fig 1).

**Fig 1.**
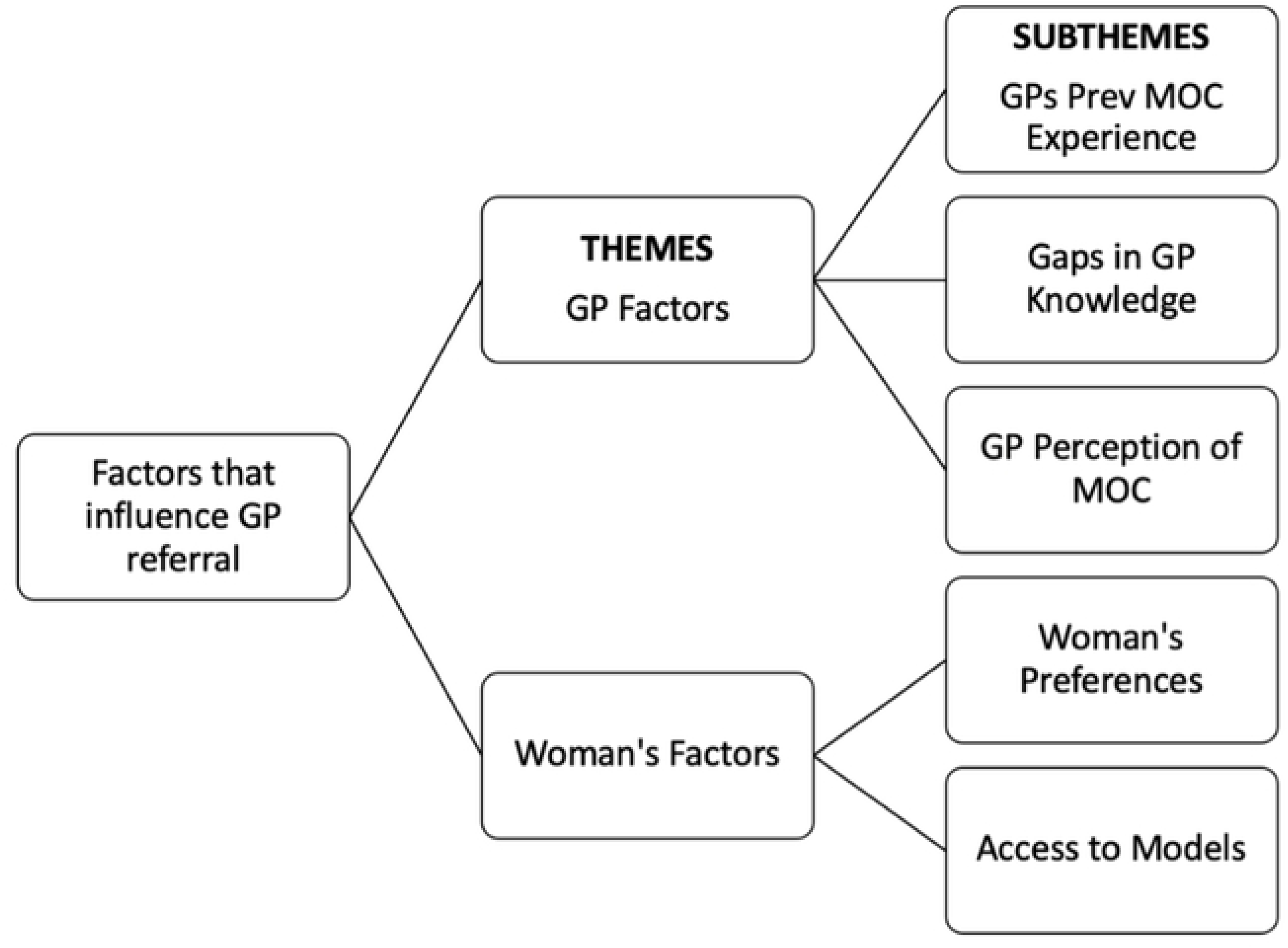
Final thematic map of the factors that influence GP referral to maternity models of care demonstrating two themes and five subthemes

## GP factors

### Gaps in GP knowledge

The participant GPs were asked about the models of care they had previously referred to and those they considered themselves to be familiar with. Overall participants had incomplete knowledge of the Australian maternity models of care and limited insight into the gaps in their knowledge base. One participant stated:

> *Well, I either go public or private because that’s all the options we have…” (P5)*

Most GPs described gaining their model of care knowledge from their work experience which predominantly occurred in a hospital setting. One GP participant said:

> *It [was] just seeing women come through the birth suite and where they’ve come from and what their journey has been and figuring out from there if it was a model or what the model was… (P7)*

The GPs were most familiar with the medical models and were less knowledgeable about midwifery-led models of care:

> *I feel like I know enough about the medical model[s] because they’re the ones that I’ve worked most in, but I feel like midwifery group practice, community midwifery programs, family birth centres we don’t get a lot of information about…” (P7)*

Of those interviewed, two GP participants mentioned their Public Health Network (PHN) when describing their model of care education and resources. Participant 3 stated that they use the PHN developed *HealthPathways* web-based portal that supports clinicians’ referrals:

> *…looking at the HealthPathways and seeing what’s actually available in my area when and I sort of do that at point of care so when a woman who comes in and requesting a referral for pregnancy I kind of open that up… (P3)*

### GP’s previous model of care experience

Previous model of care experience both personally and professionally was described as influential to the GP’s referral practice. In terms of personal birthing experience most GPs accessed private obstetric-led care. One GP said:

> *I had my first baby in the private sector, I didn’t think things went particularly well. I had my second baby as a private patient in the public sector and things went exceptionally well so I was quite impressed with the public system so that of course has influenced me (P5)*

All GPs in the cohort had professional exposure to hospital-based models of care with fewer participants describing GP obstetrics and birth centre experience. One participant said:

> *[Other GPs] might not be aware of other models of care like caseload midwifery if they haven’t had that sort of exposure (P10)*

Communication from a service was also seen as influential to the GP referral:

> *…if a service is good at communicating with me I’m more likely to refer to it as well…if that service sends me a letter with good information or lets me know if something significant has happened, then I’m more likely to want to continue to use that service (P6)*

### GP’s perception of the models of care

When asked about their rationale for making previous referrals to the maternity models of care, GPs described referring to models that they perceive positively one participant said:

> *Well, I’m happy with all of them (P4)*

GPs described referring to providers that they had confidence, faith, and trust in for example one GP stated:

> *…the most common model of care that I refer to would be delivering with general practitioners in a [regional centre] and I think that’s because I have confidence in the service (P12)*

Positive word of mouth from women was described by most GPs as a factor that increased their referrals to a service or provider stating …*you get good reports about somebody you tend to refer to them more (P12)*

On the other hand GPs said that negative feedback from women about their model of care made them less likely to make future referrals to that service.

Most interviewed GPs had an unfavourable perception of private midwifery models of care offering homebirth, one participant stated:

> *I generally don’t refer [to] that model of care ‘cause I guess …I’m more worried about their backup systems when things go wrong (P12)*

### Woman’s factors

#### Woman’s preferences

When GPs were asked about how they make their referral decisions, they overwhelmingly indicated, *…so the predominant way is through patient choice… (P10)*. This included considering the woman’s desired mode of birth or birth location.

One GP said:

> … *just talking through the pros and cons of each model, they may lean strongly towards one so they may have a preference for more midwife-led care or a preference for avoiding intervention if possible or …they’re just very keen on caesar or induction early on in the pregnancy that may influence how we make a decision…(P6)*

A patient or woman-centred philosophy was emphasised by the GP’s explaining:

> *…if you follow what they want, they tend to end up with a better outcome, sort of more happy patients and… [if] a patient is contented with their pregnancy and delivery, is more likely to be a contented mother… (P5)*

Even when questioned about their feelings towards a woman declining a referral, GPs maintained:

> *…I’d feel fine I’d respect that…she has complete capacity to make that decision about her and her baby so that would be fine…her birth her pregnancy, her choice really yeah (P11)*

#### Access to models

Access around the models of care was described by GPs as a major consideration during referral discussions.

Rural and regional GPs highlighted how women living in these areas are limited to a few models of care explaining:

> *…there’s …limited options ‘cause we live in the country…- the options are what they are… (P12)*

The maternal and fetal risk profile was described as a directing force during referral discussions. It was often referenced as a contraindication to the woman’s choice. One GP said:

> …*you know it’s their pregnancy and sometimes that doesn’t work out you know they might actually have a high-risk pregnancy and they don’t get to do what they would like to do… (P2)*

Financial and health insurance status of the woman was also highly relevant to referral discussions. GPs described discussions around out of pocket costs for women with one GP saying:

> *.. if they see me for GP shared care ’cause I’m in a private billing practice, so there will be out of pocket costs associated with every appointment that they come in to see me … when it comes to the midwifery-led models of care I sort of yeah explain what that means in terms of cost which are minimal (P1)*

## Discussion

To our knowledge, this is the first study anywhere in the world to explore factors that influence GP referral to maternity models of care. Gaps in knowledge may impact GP’s ability to offer a comprehensive range of models to women and in particular, their reduced knowledge of midwifery-led continuity of care models may mean that the benefits associated with these models such as reduced intervention and increased spontaneous vaginal birth [36], go unrealized. Given the important referring function of the GP it is likely that these referral practices may be impacting maternal and neonatal outcomes at a population health level.

Resources supplied by Primary Health Networks (PHNs) as the *HealthPathways* portal were used by two GP participants during their model of care discussions. PHNs are important to enhancing GP knowledge of the maternity models of care. PHNs assist with health service planning so as to ensure that primary care meets the community’s needs. Furthermore, PHNs address gaps in health care, acquire new services and analyse the efficacy of current services [37]. This occurs by collaboration with both health service providers and stakeholders [37]. PHN developed resources, such as the *HealthPathways* portal is used to provide local clinical management pathways and referral advice [38]. However, the portal is limited by its completeness and relevance of information. For example, the Western Australian *HealthPathways* portal does not include locally relevant model of care referral information and instead has adapted its ‘Antenatal Consult – First’ pathway from New Zealand [39]. Embedding a decision tool into the *HealthPathways* portal with the up to date and locally relevant model of care service information could provide GPs with support during model of care discussions and provide women with a broader range of model of care options.

Of the interviewed GPs, most participants described personal and work experience in the medical-led models of care and reported a heightened familiarity with those models. With most medical education and GP obstetric training occurring around medical -led- models, this medical focus appeared to impact the exposure and familiarity of GPs with non-medical led models. Moreover, GPs described referring to providers they had previously worked with, and if this predominantly occurs in the hospital setting then this reinforces referral to medical providers. Whilst there are no studies that have previously explored medical referring within the maternity context, a cross-sectional study of American primary care physicians (n=1252) demonstrated that together with other factors, past experience with a specialist was highly important to their referral practice [40]. Further, a prospective cohort study also conducted in the United States found that personal knowledge of a specialist was the most important consideration for primary care doctors when referring patients [41]. Given that most of the interviewed participants had hospital-based model of care experience, GPs may preferentially choose obstetricians working in those models of care as opposed to community-based GP obstetricians or midwives.

GPs disclosed referring to providers they perceive positively. This sentiment was echoed by Choudhry et al. stating that referral recommendations made by physicians have little or no objective basis and rather that the physicians perception of the specialist’s reputation, knowledge base and feedback from previously referred patients instead are highly valued [42]. Addressing this practice is made difficult as one needs exposure to a model to develop a positive perception of that model. However, this finding provides insights for maternity providers into aspects that are important such as communication, respectful practice and convenience for patients which impact their perception by GPs. Similar factors regarding model of care convenience for the woman and effective communication have been identified in the literature as influential to the primary care physician’s decision-making [40, 42, 43].

The woman’s choice and preferences were reported by GPs as central to the referral decision, giving insight into what they consider women view as important including birth location, mode of birth and continuity of carer. At face value, these findings may indicate GP alignment with the key intents of the national maternity strategy [15]. The reluctance demonstrated by some participants to refer to privately practising midwives in this research has been explored in previous studies [12, 44, 45]. In a national survey of n=1657 participants, narratives from women describing challenges of having to ‘fight’ with their GP to obtain a referral to a private midwife or to receive midwife only care [45]. GP reluctance or refusal to refer to midwifery models of care requires further exploration. Changes in the National Law requiring private midwives to have a collaborative arrangement with a GP are underway, it is anticipated that this will improve women’s access to private midwifery and remove the requirement for referral from a GP [46].

Finally, GPs described access around the models of care as a key consideration during model of care discussions. GPs described how rural and remote women have limited options for maternity care which is congruent with current literature that outlines how women residing in remote geographical areas experience the least antenatal care access [47]. Improving maternity care access for rural and remote women is a key objective within the national maternity strategy. Strategies such as optimising all health workforce scope, service innovations such as telehealth and outreach care as well as national pathways are critical to achieving improved access for rural and remote women [15]. The interviewed GPs also described how characteristics like maternal fetal risk profile, the financial status of the woman, and convenience, may mean that current model of care options do not meet the needs of women. Despite current risk stratification practices, there is compelling evidence regarding the benefits of continuity of midwifery care for women of all risk profiles alongside a multidisciplinary team [20, 48]. Although publicly funded models of care represent those most commonly accessed by Australian women, rising out-of-pocket costs serve as a barrier to accessing care [49]. Strategies such as funding models that support continuity of care for all women, shifting GP practice funding to support patients in need and expanding access to expert primary care through clinically activated PHNs have been discussed [15, 50, 51]. Factors of convenience such as proximity to the model of care expressed by the interviewed GPs have been highlighted as important to other doctors when undertaking a referral [40, 42, 43]. The benefit of convenient maternity models of care include reduced work and family disruption, reduced travel related safety issues, and the ability to facilitate birthing on Country, a priority for many Aboriginal women [15].

Maintaining currency of knowledge of the maternity models available within a local referring area presents a challenge for GP’s that could be met by providing online resources to support broader conversations with women and referral practices. Both national and individual jurisdictions have begun to address the gap in knowledge, examples include the Pregnancy, Birth and Baby website and the MyBaby WA App which targets new or expectant mothers but may also be used by health professionals to support a range of conversations including model of care choice [52, 53]. Education, beginning early on in medical schools, and in GPs training programs on the breadth of model of care options available is important to increase awareness and enable GPs to refer from a more empowered position. In their public health education role, GPs could further enhance women’s understanding of their options so all stakeholders can experience the benefits of consumer-demand and led access to a wider variety of models.

Within these education structures exposure to non-hospital-based models of care may provide the opportunity for GPs and training doctors to gain a sense of familiarity and enhance their knowledge of those models. This and expanding the role of midwives in the primary health sector including in primary health network (PHN) stakeholder groups may foster networks between GPs and other providers improving health service integration. Despite no current evidence evaluating the benefit of midwifery integration into primary care clinics or PHN stakeholder groups in Australia, a predominantly European study that surveyed GPs and patients across 34 countries evaluated GP outcomes and patient experiences of GPs co-located with other health professionals including midwives [54]. Of the 7183 GPs surveyed, most were co-located with nursing staff while only one in three described themselves as co-located with midwives, specialists, physiotherapists, dentists or pharmacists [54]. The analysis demonstrated that co-location with the aforementioned allied health staff or midwives resulted in improved coordination with secondary care [54]. As such, the integration of midwives into GP practices or other primary settings may confer improved coordination with secondary care however more research into the benefits of midwifery integration into the primary health sector and in PHN stakeholder groups is needed.

Finally, insights gained around women’s ability to access models of care is helpful for health system planning with a view to improve choice for women and the range of models of care available to them. Strengthening the rural workforce by training GP obstetricians, enabling scope fulfilment for midwives who have recognized expertise in primary maternity care, using telehealth and outreach services and national pathway development have been highlighted as vital to improving healthcare delivery to rural women [15, 55]. Moreover, the development of all-risk caseload midwifery models of care [20, 48], funding that supports continuity of care options [15, 50, 51] and model of care design that improves the convenience elements of maternity models [15] have the potential to better meet the needs of new and expectant parents in Australia.

## Data Availability

All relevant data are all contained within the manuscript and its Supporting Information files.

## Acknowledgments

We would like to acknowledge the general practitioners who dedicated their time to this research while still working among a global pandemic to share their experiences providing us with a deeper understanding of the factors influencing referral to maternity models of care.

## Inclusivity statement

This research uses gendered terminology. It is recognised that individuals have diverse gender identities. In this paper, pregnant persons are described with the nomenclature “woman/women”. The use of these terms is not intended to exclude those who do not identify in this way.

